# Pediatric Multi-Organ Dysfunction Syndrome: Analysis by an Untargeted “Shotgun” Lipidomic Approach Reveals Low-abundance Plasma Phospholipids and Dynamic Recovery Over 8-Day Period, a Single-Center Observational Study

**DOI:** 10.1101/2020.11.24.20237891

**Authors:** Mara L. Leimanis Laurens, Karen Ferguson, Emily Wolfrum, Brian Boville, Dominic Sanfilippo, Todd A. Lydic, Jeremy W. Prokop, Surender Rajasekaran

## Abstract

Lipids are stable molecules involved in metabolism and inflammation. We investigated the plasma lipidome for markers of severity and nutritional status in critically ill children. Children with multi-organ dysfunction syndrome (MODS) (n=24) were analyzed at three time points and cross referenced to sedation controls (n = 4) for a total of N=28. Eight of the patients with MODS, needed veno-arterial extracorporeal membrane oxygenation (VA ECMO) support to survive. Blood plasma lipid profiles were quantified by nano-electrospray (nESI), direct infusion high resolution/accurate mass spectrometry (MS), and tandem mass spectrometry (MS/MS) and compared to nutritional profiles and PEdiatric Logistic Organ Dysfunction (PELOD) scores. PELOD scores were not significantly different between MODS and ECMO cases across time-points (p = 0.66). Lipid profiling provided stratification between sedation controls and all MODS patients for lysophosphatidylserine (lysoPS) (p-value = 0.004), total phosphatidylserine (PS) (p-value = 0.015), and total ether-linked phosphatidylethanolamine (PE) (p-value = 0.03). Phospholipids in patients needing ECMO were observably closer to sedation controls than other MODS patients. Nutrition intake revealed changes in lipid profiles that corresponded to calorie and protein intake. Lipid measurement in the intensive care environment shows dynamic changes over an 8-day PICU course, suggesting novel indicators for defining critically ill children.

## 1. Introduction

It has been estimated that twenty percent of critically ill patients present to the pediatric intensive care unit (PICU) with multi-organ dysfunction syndrome (MODS) [1]. This group experiences ten times the mortality rate compared to other PICU patients [2]. In this group, there exists a smaller cohort of patients that require aggressive life support measures such as extracorporeal membrane oxygenation (ECMO) with up to 45% reported mortality [3]. The reasons for this are unclear. Understanding MODS has become increasingly important as COVID-19 patients that present with MODS experience a more protracted ICU course and suffer higher mortality than those have single system involvement [4,5].

Critically ill children experience a hypermetabolic stress response and catabolic state [6]. During this acute illness phase a rapid breakdown of adipose tissue occurs leading to multiple sequelae including chronic inflammation, hyperglycemia, and further damage to organ function by fatty infiltration [7,8]. These patients have often ceased oral intake thus being deprived of optimal nutritional support [9,10]. Critical illness by itself is characterized by mitochondrial dysfunction [11] and perturbations in β-oxidation [12] from organ failure. MODS results from a variety of triggers both infectious and non-infectious that results in a common phenotype of organ dysfunction. How lipid levels and composition vary over time and between patients with MODS alone vs. those progressing to circulatory collapse and the need for ECMO remains unknown. This knowledge could enable optimization of nutrition therapy as well as identify new targets for supporting organ function in the face of critical illness.

Unlike some biological molecules, lipids are stable and abundant, making them attractive metabolites to study in MODS patients admitted in the PICU. The role of lipids in critical illness and acute inflammation has been explored in adults [13] and children [14], and lipids are crucial for membrane formation, signaling, and metabolism [15-19]. It is well known that disruption of lipid signaling pathways and metabolism leads to inflammatory disorders [20]. This can be amplified when exogenous lipid sources from diet are altered, such as in acute illness leading to malnutrition and poor outcomes [21]. Specifically, phospholipids are known to play a role in a wide array of diseases (see review [22]). Thus, lipids likely serve as markers for tissue injury, inflammation, metabolic dysfunction, and indicators of nutritional status.

A two time-point lipidomics study focusing on single organ injury in adult patients with acute respiratory distress syndrome (ARDS) revealed 90 significantly different lipids that distinguished survivors from non-survivors [13]. We hypothesize that certain lipid classes can be used to distinguish patients with MODS that need ECMO from those that recover with just medical management accounting for both severity and nutritional status. Advances in the availability and ease of use of lipid profiling in a clinical setting allows for signatures of highly personalized lipidome profile to be developed to identify patients experiencing a severity of illness needing ECMO [23], and such a strategy has potential to be more robust than the rise or fall of a single biomarker. This study uses an unbiased untargeted lipidomics approach to enable a comprehensive study of pediatric lipid profiles, determining the molecular compositions of analytes in blood plasma using direct infusion high resolution/accurate mass spectrometry (MS) and tandem mass spectrometry (MS/MS).

## 2. Materials and Methods

### Study Population, Site and Sample Collection

Patients who were critically ill with MODS were recruited from the PICU at Helen DeVos Children’s Hospital (HDVCH), a quaternary care facility in Western Michigan, after screening for eligibility and subsequently consented. The HDVCH PICU with over 1,500 admissions per year, and over 6,000 patient days, covers a 24-bed unit. Samples were collected at up to three independent time-points: baseline, > 48 hrs, and > 7 days, as long as they remained patients of the PICU. If a patient was discharged or passed away, no further samples were collected. All patients were consented prior to study recruitment as per local IRB approval (2016-062-SH/HDVCH) [24].

Patients were referred by the attending physician on service once MODS was recognized, according to the following inclusion criteria: < 18 years of age; on vasopressors with a central line and requiring invasive mechanical ventilation for respiratory failure as per criteria established by Proulx et al. [25]. Patients presenting for routine sedation were used as controls. Patients were excluded if they had a diagnosed auto-immune disease, were considering non-interventional treatment options, had undergone cardiopulmonary bypass prior to onset of MODS, received plasmapheresis prior to ECMO initiation, or were patients of the neonatal intensive care unit at the time of consent. Blood samples were drawn in EDTA treated tubes, centrifuged, plasma was separated, and stored at −80oC. In the case of ECMO patients, samples were drawn right before going on the circuit with a median 3 days into their PICU admission (Figure 1-Study Flow Chart).

**Figure 1:**
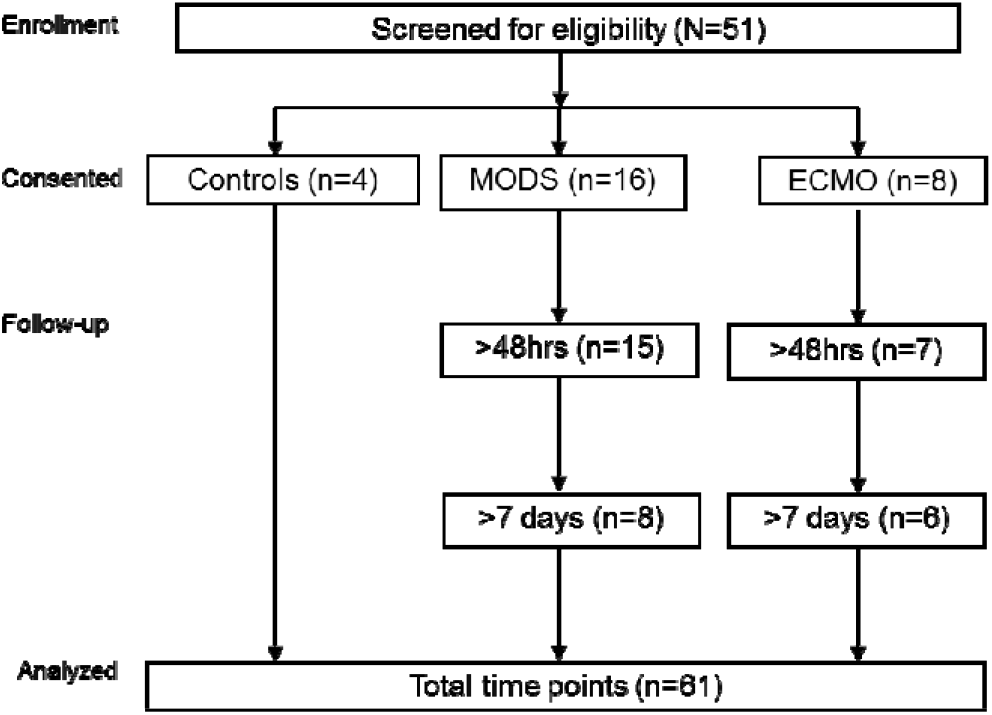
Flow diagram of the patient in the study (N=28)

### Data Collection

Basic demographic variables were extracted from the local electronic medical record (EMR). Dietary history, and mode of feeding [nil per os (NPO), per os (PO), tube feeding (TF) +/-lipids, total parenteral nutrition (TPN) +/-lipids] was extracted from the dietician’s notes in the EMR. Percent calories and protein were calculated from the resting energy expenditure [26] and daily required intake [27], and qualified according to less than or equal to 33% of needs met, 34-66%, and greater or equal to 67% nutritional needs met. Mode of respiratory support was qualified according to mechanical ventilation (MV), nasal canula (NC), or room air (RA). All study data was collected and managed using REDCap [28]. Severity of illness scores were retrieved through the Virtual Pediatric Intensive Care Unit Performance Systems (VPS, LLC, Los Angeles, CA). PELOD was included given that it is a measure of the first 10 days in the ICU. Median time to consenting patients was 2 days from ICU admission, and Pediatric Index of Mortality 2 (PIM2) and Pediatric Risk of Mortality III (PRISMIII) were both calculated during the first hours of ICU admission only, therefore not included in our study analysis.

### Blood Plasma Lipidomics Method

Blood samples collected in EDTA-treated tubes were immediately placed on ice and then spun at 4oC (once for 15 minutes at 1500rpm; a second spin for 10 minutes at 10,000rpm), plasma was harvested and frozen to −20oC, and −80oC for long-term. Lipidome profiles were determined from thawed plasma, subjected to lipid extraction with acetone, methanol, and acetonitrile [29], followed by analysis by nano-electrospray direct infusion high resolution/accurate mass spectrometry and tandem mass spectrometry utilizing an LTQ-Orbitrap Velos mass spectrometer (Thermo Scientific, Waltham, MA) with the FT analyzer operating at 100,000 resolving power over two minutes. An Advion Nanomate Triversa (Advion Biosciences, Ithaca, NY) served as the nano-electrospray source with high-throughput autosampler. To verify lipid identities, Higher-Energy Collisional Dissociation was utilized (at 100,000 resolving power). Lipidome analysis provided untargeted assessment across all classes of glycerolipids (GC) (including mono-, di- and triglycerides), phospholipids (PS), lyso-phospholipids (lysoPL), sphingolipids (SP), sterols, non-esterified fatty acids (NEFA’s) and fatty acids (FAs). Additional species analysis was completed on phospholipids, triacylglycerides (TGs), diacylglycerides (DGs), cholesterol (chol), and sphingomyelins (SMs). Global lipidome analysis provided untargeted analysis with detection limits of 0.01 nM to 10 nM. Each sample was run twice (positive ion and negative ion analysis) and data was combined. Di-myristoyl phosphatidylcholine served as an internal standard. Peak finding and quantification for global lipidomics and targeted lipid mediators was performed with Lipid Mass Spectrum Analysis (LIMSA) version 1.0 software [30] and MAVEN software [31]. Large blood volumes from this patients population was challenging to obtain, therefore plasma volumes were small (∼0.050-0.075 ml total) and higher abundance lipids (∼10-1-101) were targeted [32].

### Analysis

Percent data were transformed before being analyzed with a beta regression from the R [33] package betareg [34]. Total normalized ion values were log-transformed and analyzed using generalized linear regression models (glm) in R [35]. All regression models were adjusted for age and sex. Contrasts between treatment groups (sick vs. sedation) were conducted using R package emmeans [36]. P-values from the regression models have been corrected for multiple testing via the FDR method. Additional statistical tests that were performed include Welch’s t-tets, independent t-tests, and Wilcoxon Rank Sum tests. The p-values from these tests were not multiple testing corrected. Lipid analysis was done on all major lipid classes (as stated above). In this paper we focused on phospholips, given they are heavily influenced by exogenous dietary sources.

## 3. Results

### Study population

Basic characteristics revealed a majority of male (60.6%) Caucasian (57.1%) patients (Table 1). Ages ranged from neonates to adolescents (0.14 - 202 months) with median values of 94.25 months for control patients (range 28.0-122.5), 114.50 months for MODS (range 0.14-202) and 3.5 months for ECMO patients (range 0.5-202), age ranges for both patient groups were similar. Total hospital length of stay (HLOS) ranged between 5-377 days, and total PICU LOS ranged from 3-79 days; ECMO patients spent almost two times as long in the PICU. The majority of patients had a diagnosis of bronchiolitis/pneumonia (7/24; 29%), or sepsis (7/24; 29%). There were two mortalities at 1 years’ time, both were patients requiring ECMO. All MODS patients at baseline were mechanically ventilated, had their lungs and heart compromised, and were administered inotropes to support cardiovasular function. More than half of the participants exhibited renal dysfunction (n=15; 54%), including 7 MODS and all 8 ECMO patients (odds ratio: 6.01, p-value=0.01). Other organs affected included liver (n=8; 28.6%; 4 MODS; 4 ECMO) and brain (n=6; 21.4%; 4 MODS; 2 ECMO) in approximately one-quarter. Severity of illness scores Pediatric Logistic Organ Dysfunction-2 score (PELOD) were not significantly different between MODS and ECMO groups across time-points (Global F-stat, p=0.66). This suggests that common metrics of severity do not reflect need for ECMO further suggesting need for additional clinical stratifiers, such as assessing nutrition or lipidomics.

**Table 1.**
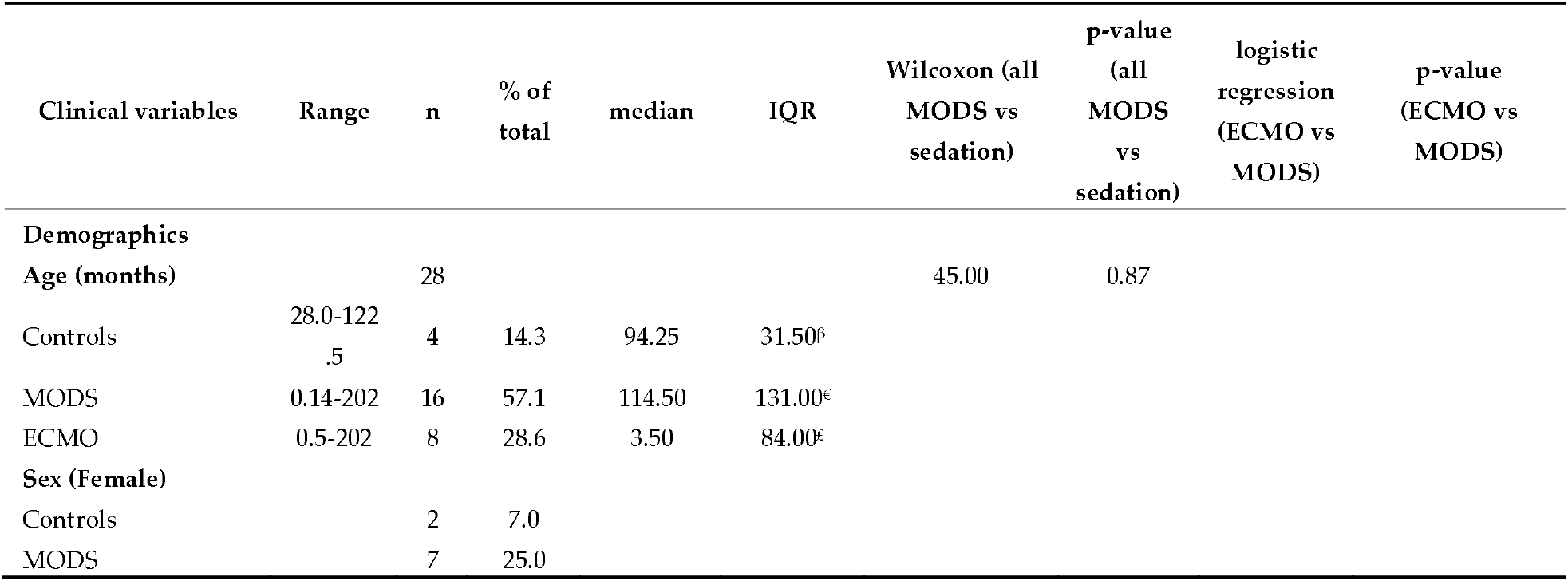
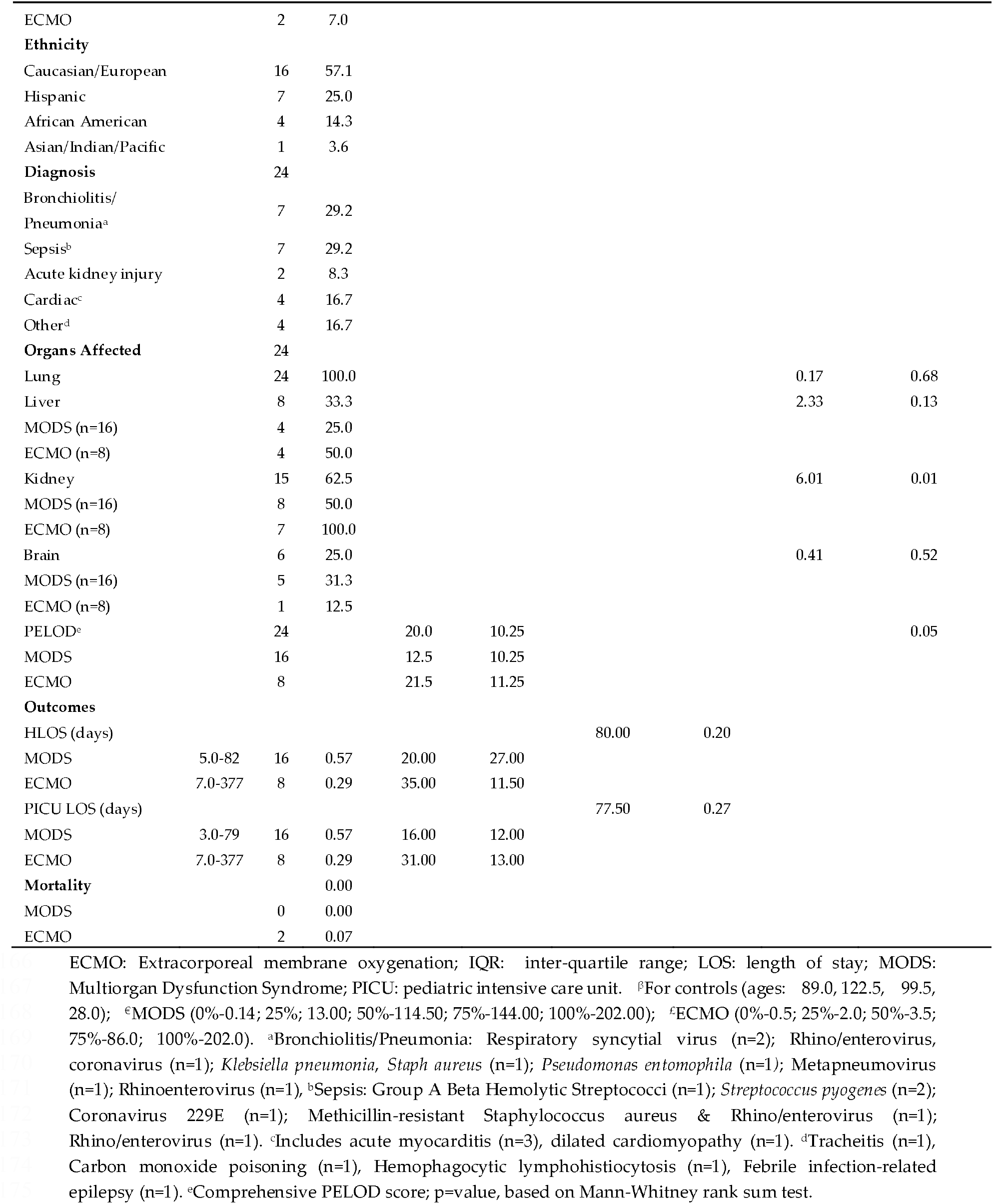
Patient characteristics of MODS/ECMO Study 2016-2018 (N=28)

### Baseline Lipid Profiles of Critically ill Children Compared to Controls

In order to determine the source of differences in lipid classes at baseline, when most patients (67%) were NPO, we examined absolute normalized abundance (per milliliter of plasma, Figure 2). Lipids in MODS patients (ECMO & MODS combined) differed significantly at baseline from sedation control samples after correcting for multiple testing based on generalized linear models adjusted for age and sex for three classes of phospholipids: lysophosphatidylserine (lysoPS) (P – value = 0.004), total phosphatidylserine (PS) (P – value = 0.015), and total ether-linked phosphatidylethanolamine (PE) (P-value = 0.03) (Figure 2A). Point estimates and confidence intervals of the differences are presented in Figure 2B. The heatmap revealed relative baseline levels for all lipid classes analysed including NEFAs, glycerolipids and TGs (Figure 2C).

**Figure 2:**
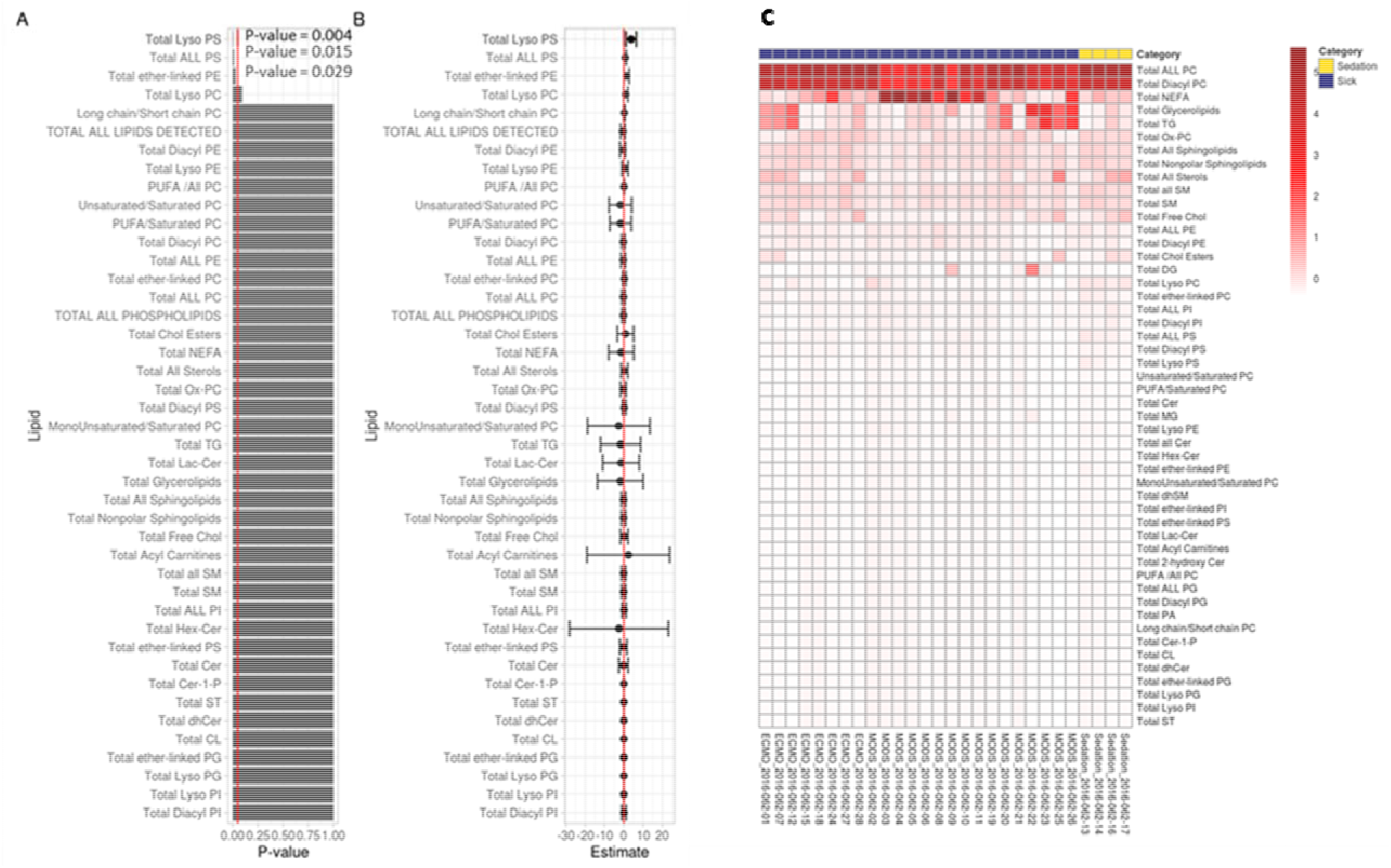
Total Baseline Lipids Based on Absolute Normalized Ion Abundances per ml of Plasma **A-**Bar chart of p-values from regression output; red line indicates 0.05. Any p-value that crosses 0.05 is not statistically significant. **B-**Points in B represent the mean log(fold-change) in odds between sick and sedation groups. The error bars in B, represent the upper and lower bounds of the false-coverage for the estimate. 95% false coverage intervals of the mean log (fold-change) in odds; red line indicates 0. Any confidence interval that crosses 0 is not statistically significant. **C-**Scaled values of total lipids based on absolute normalized ion abundances per ml of plasma.

NEFA levels were further explored and found to be significantly associated with sex. However, after adjusting for sex, none of the experimental groups were significantly associated with NEFA levels at any time point.

To serve as an additional internal control, total plasma serine (O-acetyl-L-serine) levels were compared between the three groups, from a metabolite data-set on the same patient cohort (to be presented in a separate report). Mean values were not significantly different (sedation vs MODS; p = 0.8281; sedation vs. ECMO; p = 0.3348) based on independent t-test. This may imply that in spite of plasma serine levels, the PS and lysoPS values differ for all MODS patients due to other metabolic drivers.

### Mode of Feeding and Nutritional Intake

With detailed notes of nutrition for control and MODS patients, the data can be used to investigate feeding and nutritional intake to elucidate biases in the cohort and to qualitatively control for intake into the lipidomic analyses below. Mode of feeding and percent caloric and protein intake were reviewed for each patient (Table 2). Sedation controls were NPO (nil per os, nothing by mouth) for eight hours prior to procedure. At baseline, 16/24 (67%) of all combined MODS (MODS + MODS who required ECMO) patients were NPO, which decreased (to 7%) by day 8. Baseline caloric intake was less than 33% of recommended intake for most patients, which gradually improved over time. A similar profile was reflected in the percent total protein. ECMO patients by eight days were achieving higher percent calories and protein than their MODS counterparts (over 34% goal reached): ECMO (n=5; 83%) vs. MODS (n=4; 50%) and ECMO (n=6; 100%) vs. MODS (n=4; 50%), respectively. There was no evidence of a statistical difference in the percentages of protein or total calories when tested with with Fisher’s exact test (p = 0.085; p = 0.30). All patients were experiencing a deficit nutritionally, however ECMO patients were reaching superior nutritional intake, which may be due to their being more hemodynamically stable because of the circulatory support, better facilitating their caloric need. Of further importance is how this exogenous nutritional intake influences blood plasma lipid profiles for patients over time. Based on this result, we might anticipate that ECMO patients fare better than MODS in terms of overall lipid profiles.

**Table 2.**
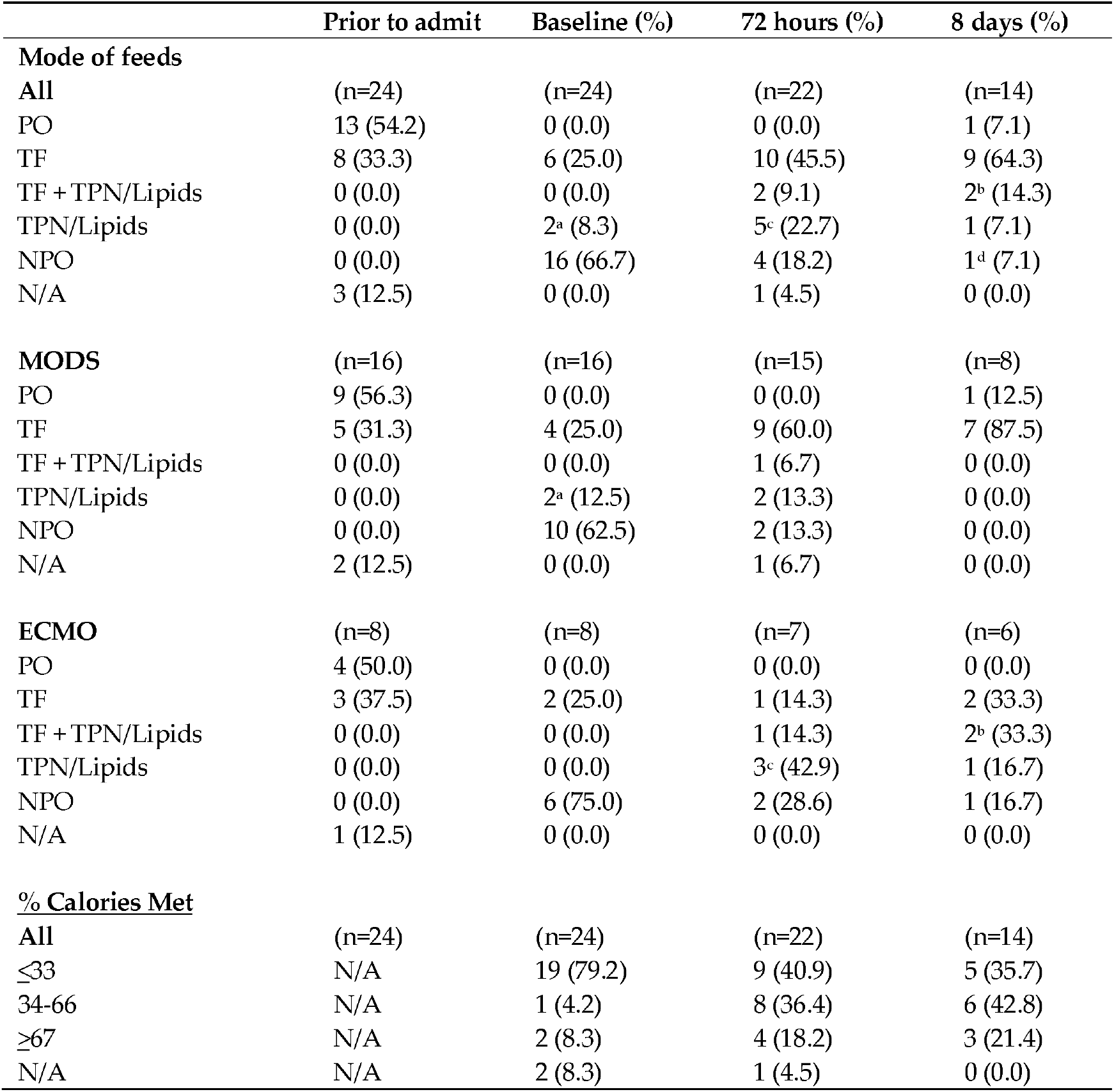
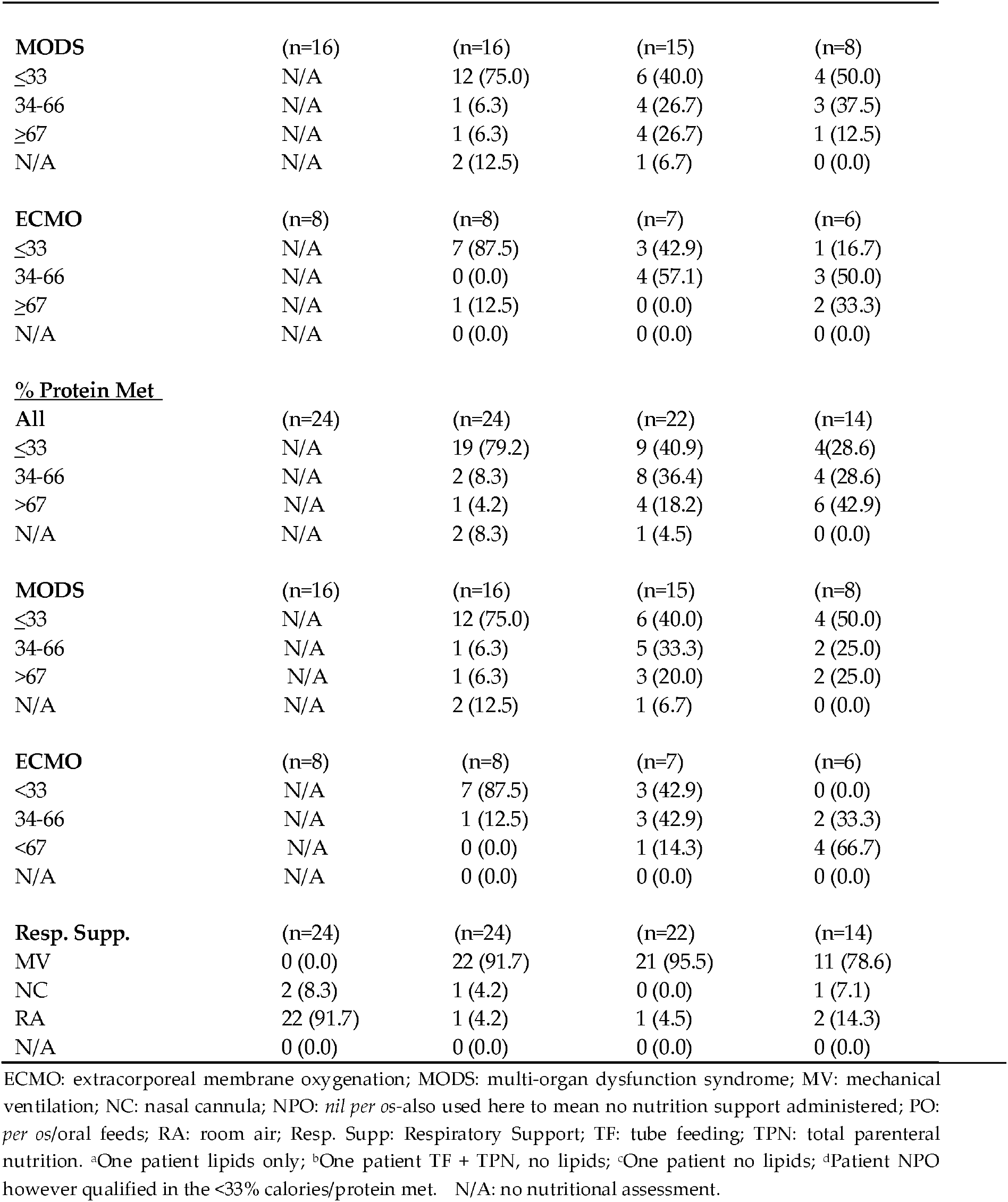
Patient Nutrition for MODS/ECMO Study 2016-2018 (N=24)

### Phospholipids of Critically ill Children Over Three Time Points

Surprisingly, for MODS patients that needed ECMO, their distribution of phospholipids appear more similar to patients coming to the hospital for same day sedation instead of looking like that of other MODS patients (Figure 3). Phospholipid levels appeared to increase with percent nutritional (caloric and protein) needs being met (Table 2). A total of five patients (n=2 MODS; n=3 ECMO) over seven time points received IV lipids as a portion of their nutritional regimen. Two patients received lipids at more than one time-point, both with slight increases in their plasma phospholipid levels.

**Figure 3:**
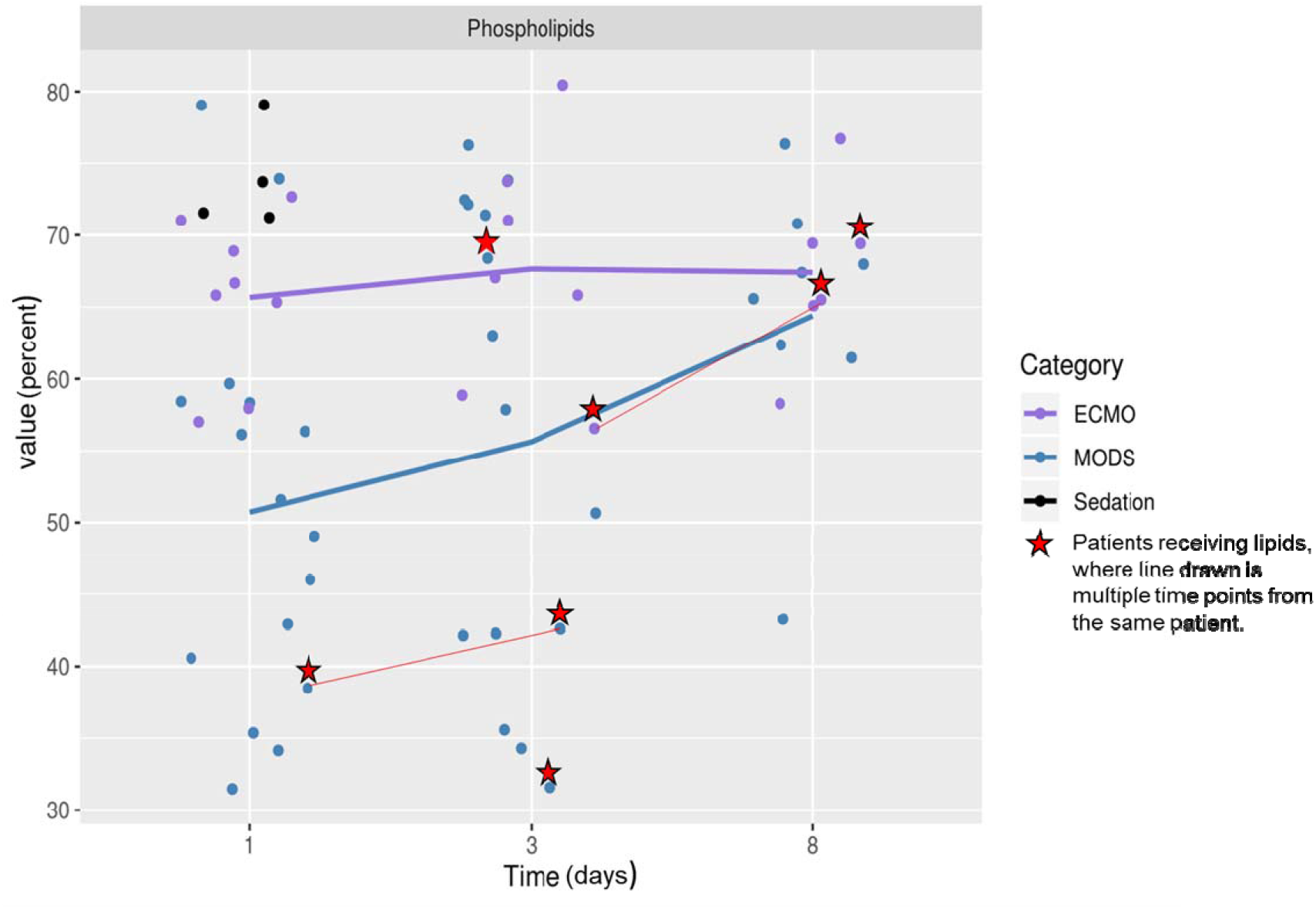
Time course for lipids over 8 days.

These differences may not be based on nutrition intake alone, given most patients were NPO (67%) at baseline, and receiving less than 33% of needed intake (79%), suggesting other metabolite drivers at play.

In summary, we see down-regulated phospholipid levels including sub-classes (lyso PS, PS, PE) at baseline for patients with MODS, with increases by day 8, except for those with MODS that needed ECMO (Figure 2, 3). We observe increases in phospholipids that correspond to nutritional intake patients received.

## 4. Discussion

Patients with MODS present with dyslipidemia at first detection of organ pathology based on our lipidomic profiles presented in this report. Patients with MODS are compromised at the time of baseline, undergoing membrane disruption/remodeling, catabolic state, canabalism, and lipid dysregulation [37], resulting in decreased phospholipids as has been previously reported for pneumonia [38] and septic patients [39]. In contrast to the MODS patients, ECMO patients have phospholipid profiles that are observedly different at baseline and throughout the eight-day course. Some reports state that phospholipids from endogenous sources such as the hepatobiliary system [40] are involved with signaling of the innate immune response [41]. In the event of multi-organ dysfunction, with liver involvement (as we see in half of our ECMO patients), perhaps this is not entirely surprising. We suspect that this observed difference in ECMO patients may be tied to severe cardiac dysfunction with an inability to compensate metabolically. Generally, risk factors for this sub-population may include pre-existing conditions such un-diagnosed conditions, metabolic syndrome or other organ pathologies, which has been seen in current COVID-19 cases [42].

Recent studies in COVID 19 infections have found patients that suffer from multi-organ involvement are more prone to mortality [43]. A subclass of PS, ether-linked phosphatidylester were found to be among the lowest-abundance lipids in COVID patients. An untargeted lipidomics approach was adopted for COVID-19 patients which revealed lower levels of certain classes of lipids which persisted even with a regular diet and discharge home [44]. This finding is indicative of ongoing metabolic disruption, post-ICU admission.

Specifically, a sub-class of lyso lipids known as -Lyso phosphatidylserine (LysoPS) has immune function [45,46] through enhanced clearance of neutrophils [47]. Synthesized by neutrophils [48-50], LysoPS enhances efferocytosis (“to carry to the grave” [51]) of neutrophils by macrophages [52] during acute inflammation [53,54]. Neutrophil levels were lower and more like controls in patients that needed ECMO [55] and other studies have shown that neutrophil levels drop in the sickest patients as bone marrow activity becomes suppressed. This may lead to the lower levels of LysoPS.

Over the past decades, the externalization of PS has been linked with efferocytosis, and characterized as the ultimate “eat-me” signal, is an evolutionarily conserved immunosuppressive signal, which prevents local and systemic immune activation [56]. This externalization of PS has been shown to be exploited by viruses, microorganisms, and parasites to promote infection [56]. This could link PS levels seen in our cohort to infection in our cohort as many of our patients presented with bronchiolitis, pneumonia and sepsis, two of which had Coronavirus (OC43 and HKU1). Many of the patients in this study had viral illness as a trigger or in addition to MODS, as is typical in the PICUs across the country.

Untargeted lipidomic research done previously in adult patients (n=30) with acute respiratory distress syndrome (ARDS) using a shot-gun lipidomics approach revealed 90 significantly different lipids that distinguished survivors from non-survivors[13]. Similarly, in children it has been shown that profiling metabolites for septic shock and systemic inflammatory response syndrome (SIRS) can yield markers of mortality [23].

This study shows that phospholipids in ECMO patients observedly grouped closer to the sedation patients. In all these patients, the lipidome is reflective of an amalgam of multiple pathways that include the inflammatory cascade, immune dysfunction, severity of illness and nutritional status, all of which could combine to effect clinical course. Age was accounted for in the models (given ECMO patients were younger than the MODS), as was suspected that immature immunity or active suppression of the immune system and a confounder [57]. ECMO patients seem to have their ability to modify their lipidome impeded as the concentrations of major lipid classes are closer to sedation controls.

## 5. Conclusions

In this prospective observational study we studied several lipid classes at multiple time-points and identified that phospholipids showed differences between all MODS and sedation, and MODS vs. MODS needing ECMO using a shotgun lipidomics approach. Future work may include a targeted lipidomic approach on phospholipids. Patient lipid profiles can be partially modified with nutritional intervention.

## Data Availability

Data will be made available following peer-review, and manuscript acceptance. Available on request.

## Supplementary Materials

N/A.

## Author Contributions

SR and ML conceived of the original study design, screened, recruited and collected all samples. ML and EW were the statistical and bioinformatics support. KF reviewed all of the nutrition information for individual patients. Peak findings, annotation, and quantification for lipidomics were performed by TL. SR, DS, ML, EW, BB, JP, KF and TL all contributed to the writing and editing of the manuscript.

## Acknowledgments

The authors would like to thank the PICU staff at Helen DeVos Children’s Hospital for their support in the completion of this study and various contributions, and Brittany Essenmacher, Dr. Jocelyn Grunwell and Dr. Eric Kort for their critical review of the final manuscript.

